# Waiting time for scheduled outpatient specialist consultations by access pathway in public hospitals in Ecuador

**DOI:** 10.64898/2026.05.04.26352408

**Authors:** Marcelo Armijos Briones, Emilia Diaz Cercado, Oscar Marcillo-Toala, Patricia Estefanía Ayala Aguirre, Pablo Lenin Benitez Sellan, Antonio Lanata-Flores, Nicole Armijos Bazurto

## Abstract

**Objective:** To quantify waiting time in days for scheduled outpatient specialist consultations and to compare waiting time between standardized and non-standardized access pathways in Ecuadorian public hospitals.

**Methods:** We analyzed hospital-based survey data from Ecuadorian public hospitals, restricted to adults attending a scheduled outpatient specialist consultation (n = 4,436). Emergency care, unscheduled urgent visits, procedures, and follow-up visits were excluded by design. Access pathway was classified from participants’ self-report as standardized (institutional or system-based) or non-standardized (informal or non-system-based). Waiting time, defined as the number of days between obtaining the appointment and attending the consultation, was compared using the Mann–Whitney U test. Sociodemographic correlates of non-standardized access were examined using adjusted logistic regression, and adjusted median differences were estimated using quantile regression (τ = 0.50). Analyses were stratified into direct-access specialties and referral-required specialties.

**Results:** Non-standardized access was associated with shorter waiting times than standardized access. In adjusted median regression, non-standardized access was associated with a 3.2-day shorter median waiting time (95% CI −4.6 to −1.8). The difference was larger in direct-access specialties (−15.0 days, 95% CI −15.0 to −6.0) than in referral-required specialties (−5.0 days, 95% CI −5.0 to 0.0).

**Conclusion:** Among patients who attended a scheduled outpatient specialist consultation in Ecuadorian public hospitals, non-standardized access was associated with shorter waiting times, particularly in direct-access specialties. These findings suggest that, within routine outpatient care, parallel access pathways may shape timeliness and warrant greater transparency in appointment allocation and referral coordination.

## Introduction

Timely access to specialist care is a core dimension of effective health system performance and quality. Delays might result in avoidable suffering, disease progression, and inefficient use of resources [1]. Conceptual frameworks emphasized that access to care was not only determined by service availability, but also how the population could identify needs, seek care, reach services, and obtain appropriate care without delay [2]. Waiting time is often used as a non-monetary rationing mechanism in publicly financed systems, however the burden of waiting is not evenly distributed across socioeconomic groups [3–5]. Evidence from different high-income settings showed that income and education have been related with shorter waiting times for specialist or planned care, challenging the assumption that waiting lists automatically delivered “*equal treatment for equal need*” [4,6,7].

In Latin America, health-system reforms have expanded coverage and benefits, yet fragmentation and persistent inequalities continue to shape access to specialist care. Delays in referral pathways, weak coordination between levels of care, and administrative barriers remain important challenges in public healthcare networks. However, patient-level empirical evidence on how standardized and non-standardized access pathways relate to waiting time for outpatient specialist consultations remains limited in the region. Evidence suggests that incomplete referral information, limited feedback from specialists, and inadequate communication tools are the most common shortcomings in public health networks, which may contribute to delays and inefficiencies in the outpatient referral process [8]. Recent regional analyses have highlighted persistent operational barriers affecting timely access to care in Latin America and the Caribbean [9].

A key reason this gap matters is that “access” is not only defined by standard rules. An classic work conceptualized access as the fit between patients and health systems across dimensions including availability, accessibility, and accommodation [2,10]. When standard (system-based) pathways are being constrained by capacity, administrative complexity, or weak coordination, patients might seek non-standardized (non-system-based) routes, through personal networks or other mechanisms, to obtain appointments with shorter intervals. If such non-standardized routes systematically reduced waiting time for some individuals, then timeliness might become partly determined by social connections rather than clinical needs, widening inequities even within a publicly financed system.

In Ecuador, the Comprehensive Health Care Model (Modelo de Atención Integral de Salud, MAIS) organizes care across levels of complexity and formally relies on a referral and counter-referral subsystem [11]. Within this framework, outpatient specialist care in public hospitals offers an informative setting to examine waiting time and access pathways, particularly because some specialties may be accessed in routine practice with weaker referral constraints than others. The aim of the present study was to quantify waiting time in days for scheduled outpatient specialist consultations and to compare waiting time between standardized and non-standardized access pathways. We also examined sociodemographic correlates of non-standardized access and explored whether the association between access pathway and waiting time differed between direct-access and referral-required specialties.

## Methods

### Study design and setting

This was a cross-sectional observational study based on a hospital survey conducted in public hospitals in Ecuador. The study design, site selection, field procedures, and questionnaire development were described in a published protocol [12]. The present manuscript reports a focused analysis of waiting time and access pathways for scheduled outpatient specialist consultations.

### Participants and eligibility

Participants were interviewed outside public hospitals before entering the outpatient specialist area. Eligible participants were adults attending a scheduled outpatient specialist consultation in the public hospital network. Emergency care, unscheduled urgent visits, appointments for procedures, and follow-up visits with the same specialist were excluded by design because the present analysis focused on first scheduled outpatient specialist consultations.

### Data collection

After informed consent, trained interviewers administered the questionnaire verbally and recorded responses in a structured Google Forms survey, which uploaded the information to a centralized database in real time. Interviewers were deployed across the country, except in the Galápagos Islands for logistical reasons. Data collection took place outside hospitals because institutional authorization for in-hospital recruitment was not available; as interviews were conducted in public space before outpatient entry, this procedure did not require additional authorization from the Ministry of Public Health. Data collection outside public hospitals began in July 2024 and ended in December of the same year.

### Exposure definition

Standardized access was defined as obtaining the appointment through the institutional referral pathway of the public health system. According to the original study protocol, this included referral from a doctor at a health centre, referral from a specialist at this or another hospital, and appointment scheduling at the hospital or by telephone. Non-standardized access was defined as obtaining the appointment through non-system-based mechanisms, including through a friend or family member or through a person who was not a friend or family member.

### Outcome definition

Waiting time was defined as the self-reported number of days between the date on which the outpatient specialist appointment was obtained and the date on which that scheduled consultation was attended.

### Covariates

Self-reported covariates included age, sex, education level, geographic region (Sierra, Coast, Amazon), ethnicity, area of residence (urban or rural), and household income. Income was recorded numerically and categorized into sample-based quintiles (Q1 lowest to Q5 highest).

### Specialty stratification

For the stratified analyses, specialties were grouped a priori according to the referral requirement in routine practice. Dentistry, General Medicine, Family Medicine, and Psychology were classified as direct-access specialties, since these specialties are directly accessible in hospitals; that is, a referral is not required to access them. All other specialties were classified as referral-requiring specialties.

### Statistical analysis

Participant characteristics were summarized overall and by access pathway. Continuous variables were described using means and standard deviations when approximately symmetric, and medians with interquartile ranges (IQR) for skewed distributions. Categorical variables were summarized using counts and percentages.

Waiting time distributions were compared between access pathways using the Mann–Whitney U test, consistent with the non-normal distribution of waiting time.

To examine sociodemographic correlates of non-standardized access, we fitted a binary logistic regression model with non-standardized access as the dependent variable and reported crude and adjusted odds ratios (ORs) with 95% confidence intervals (CIs). The adjusted model included age, sex, education, geographic region, ethnicity, household income quintile, and area of residence.

To estimate the adjusted association between access pathway and waiting time in absolute days while accommodating right-skewness, we applied quantile regression at τ = 0.50 and reported the adjusted median difference (non-standardized minus standardized) with 95% CI.

To assess whether the association between access pathway and waiting time differed by specialty referral requirement, analyses were stratified into direct-access and referral-required specialties. Within each stratum, we report medians (IQR) by access pathway and the median difference (non-standardized minus standardized). Confidence intervals for stratum-specific median differences were estimated using bootstrap resampling.

Analyses were restricted to complete cases for variables required in each model.

Because this study focused exclusively on scheduled outpatient specialist consultations, emergency presentations and unscheduled urgent care were excluded by design. In the Ecuadorian public system, patients with acute urgent conditions are generally directed to emergency services rather than managed through routine outpatient scheduling. However, some scheduled outpatient consultations may still differ in perceived clinical priority, and this factor was not directly measured.

### Ethics

This study was approved by the Ethics Committee for Research in Human Subjects of the Portoviejo Higher Technological University Institute (CEISH-ITSUP) under code 1718079732. The committee is registered with the Ministry of Public Health of Ecuador and with the Office for Human Research Protections under number IRB00014260. All participants provided informed consent before the interview. No directly identifiable personal information was collected.

## Results

The analytic sample comprised 4,436 adults who attended a scheduled outpatient specialist consultation in public hospitals in Ecuador. Overall, 1,431 participants (32.3%) reported standardized access and 3,005 (67.7%) reported non-standardized access. The mean age was 43·22 years (SD 15·38), and women accounted for 63·5% of the sample. Most respondents reported secondary education (51·5%), followed by primary education (26·9%), and third-level education (17·7%), while 0·5% reported fourth-level education. Most participants lived in urban areas (79·0%). Regarding geographic region, 45·3% resided in the Andean highlands (Sierra), 35·4% in the Coastal region (Costa), and 19·4% in the Amazon region (Amazonian). Most respondents self-identified as Mestizo (87·9%), while 4·7% identified as Indigenous. Because waiting time was right-skewed, it was summarized using medians and interquartile ranges; the overall median waiting time was 30 days (IQR 15–45). Median waiting time was 30 days (IQR 15–45) among participants reporting standardized access and 25 days (IQR 12–40) among those reporting non-standardized access.

**Table 1.** Sociodemographic characteristics and waiting times of study participants, by access pathway. Ecuador, 2025.

| Variable | Total (n = 4436) | Standardised (n = 1431) | Non-standardised (n = 3005) |
| --- | --- | --- | --- |
| Age, mean ± SD | 43.22 ± 15.38 | 43.93 ± 16.32 | 42.89 ± 14.90 |
| Sex: Female | 2818 (63.5) | 951 (66.5) | 1867 (62.1) |
| Sex: Male | 1618 (36.5) | 480 (33.5) | 1138 (37.9) |
| Education: None | 150 (3.4) | 52 (3.6) | 98 (3.3) |
| Education: Primary | 1195 (26.9) | 365 (25.5) | 830 (27.6) |
| Education: Secondary | 2286 (51.5) | 733 (51.2) | 1553 (51.7) |
| Education: Third level | 785 (17.7) | 270 (18.9) | 515 (17.1) |
| Education: Fourth level | 20 (0.5) | 11 (0.8) | 9 (0.3) |
| Area of residence: Urban | 3503 (79.0) | 1079 (75.4) | 2424 (80.7) |
| Area of residence: Rural | 933 (21.0) | 352 (24.6) | 581 (19.3) |
| Geographic region: Andean highlands | 2008 (45.3) | 654 (45.7) | 1354 (45.1) |
| Geographic region: Coastal | 1569 (35.4) | 532 (37.2) | 1037 (34.5) |
| Geographic region: Amazon | 859 (19.4) | 245 (17.1) | 614 (20.4) |
| Ethnicity: Mestizo | 3901 (87.9) | 1238 (86.5) | 2663 (88.6) |
| Ethnicity: Indigenous | 209 (4.7) | 81 (5.7) | 128 (4.3) |
| Ethnicity: Afro-Ecuadorian | 161 (3.6) | 71 (5.0) | 90 (3.0) |
| Ethnicity: Montubio | 103 (2.3) | 19 (1.3) | 84 (2.8) |
| Ethnicity: White | 62 (1.4) | 22 (1.5) | 40 (1.3) |
| Household income quintile: Q1 | 888 (20.0) | 229 (16.0) | 659 (21.9) |
| Household income quintile: Q2 | 887 (20.0) | 244 (17.1) | 643 (21.4) |
| Household income quintile: Q3 | 887 (20.0) | 280 (19.6) | 607 (20.2) |
| Household income quintile: Q4 | 887 (20.0) | 393 (27.5) | 494 (16.4) |
| Household income quintile: Q5 | 887 (20.0) | 285 (19.9) | 602 (20.0) |
| Waiting time (days), median (IQR) | 30.0 (15.0–45.0) | 30.0 (15.0–45.0) | 25.0 (12.0–40.0) |

Waiting time to specialist appointments differed by access type. As shown in Figure 1, the distribution of waiting time was right skewed in both groups; therefore, results were summarized using medians and interquartile ranges. Participants with standardized access had a median waiting time of 30 days (IQR 15–45), whereas those reporting non-standardized access had a median waiting time of 25 days (IQR 12–40). The difference was statistically significant (Mann– Whitney U = 2,433,451; p < 0·001), with a small effect size (rank-biserial r = −0·132), indicating shorter waiting times among participants who reported non-standardized access. For visualization purposes, the y-axis was truncated at 200 days; three observations above this threshold were not displayed, without affecting the statistical comparison, which was conducted using the full dataset.

**Fig 1:**
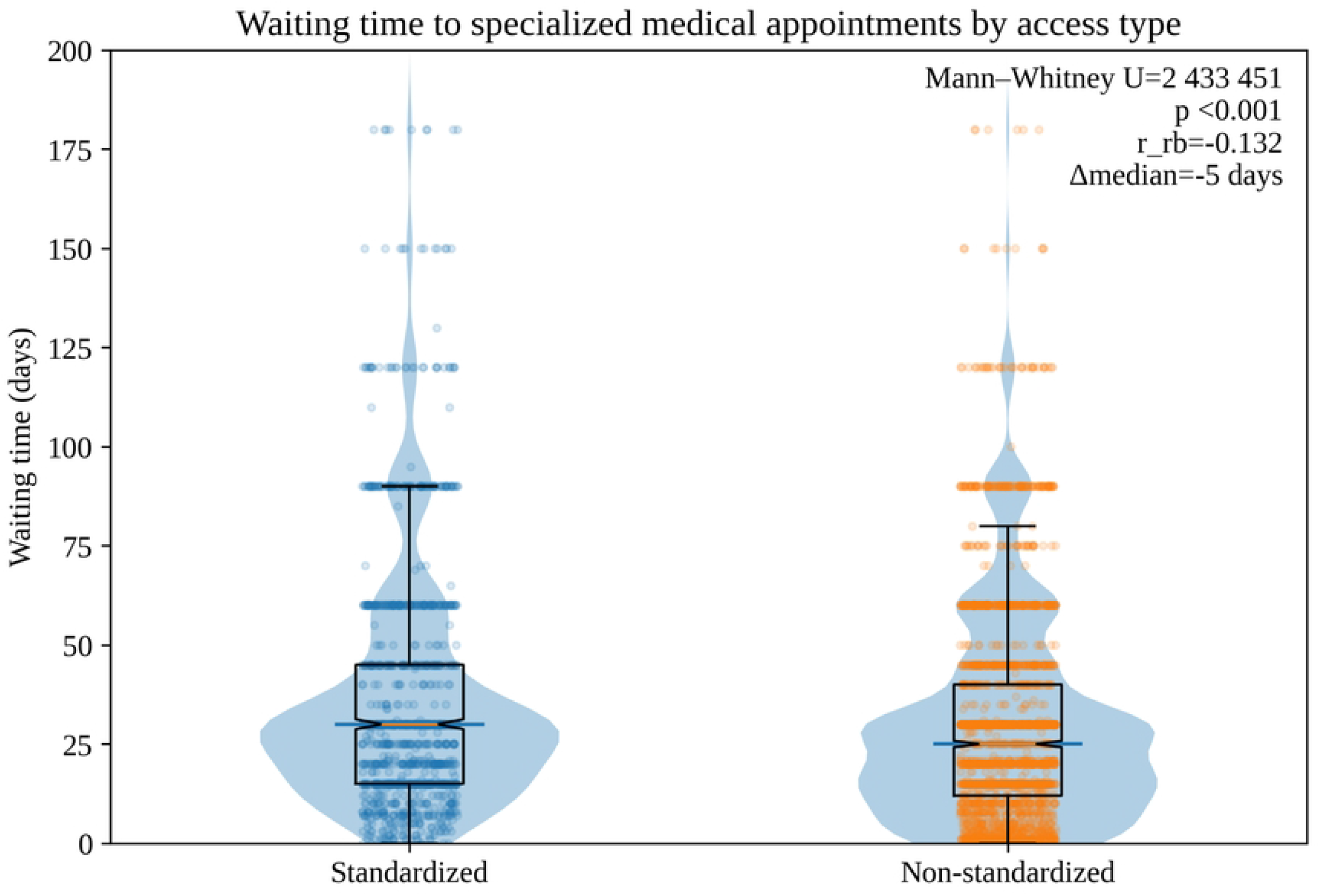
Waiting time for specialist appointments by access type (standard vs non-standard).

Figure 1 shows the distribution of waiting time in days. The inset shows the results of the Mann-Whitney U test and the rank-biserial effect size. For ease of visualization, the Y-axis was truncated at 200 days; the three observations exceeding this threshold are not shown.

In crude analyses, several sociodemographic factors were associated with reporting non-standardized access to specialist appointments (Table 2). In the multivariable model adjusting for sex, age, education, geographic region, ethnicity, household income quintile, and area of residence, female participants had lower odds of non-standardized access compared with males (aOR 0·79, 95% CI 0·69–0·91), and rural residents had lower odds compared with urban residents (aOR 0·67, 95% CI 0·57–0·80). Geographic region also showed differences after adjustment, with participants living in the Amazonian region having higher odds of non-standardized access compared with those in the Sierra (aOR 1·21, 95% CI 1·01–1·44), whereas the Costa region did not differ from the Sierra. Socioeconomic gradients were evident: compared with the highest income quintile, the lowest (Q1) and Q2 quintiles had higher odds of non-standardized access (Q1 aOR 1·55, 95% CI 1·23–1·96; Q2 aOR 1·31, 95% CI 1·05–1·63), while Q4 had lower odds (aOR 0·61, 95% CI 0·50–0·75). Age showed no clear association with non-standardized access after adjustment (aOR 1·00, 95% CI 0·99–1·00), and adjusted estimates for education and ethnicity were broadly consistent with no strong differences.

**Table 2.** Crude and adjusted odds ratios for non-standardised access to specialist appointments. Ecuador, 2025.

| Variable | Crude OR (95% CI) | Adjusted OR (95% CI) |
| --- | --- | --- |
| Sex: Male (reference) | 1·00 (ref) | 1·00 (ref) |
| Sex: Female | 0·83 (0·73–0·95) | 0·79 (0·69–0·91) |
| Education level: Secondary (reference) | 1·00 (ref) | 1·00 (ref) |
| Education level: None | 0·89 (0·63–1·26) | 0·90 (0·62–1·31) |
| Education level: Primary | 1·07 (0·92–1·25) | 1·02 (0·87–1·20) |
| Education level: Third level | 0·90 (0·76–1·07) | 0·95 (0·79–1·14) |
| Education level: Fourth level | 0·39 (0·16–0·94) | 0·41 (0·17–1·00) |
| Geographic region: Sierra (reference) | 1·00 (ref) | 1·00 (ref) |
| Geographic region: Costa | 0·94 (0·82–1·08) | 0·89 (0·77–1·03) |
| Geographic region: Amazonian | 1·21 (1·02–1·44) | 1·21 (1·01–1·44) |
| Ethnicity: Mestizo (reference) | 1·00 (ref) | 1·00 (ref) |
| Ethnicity: Indigenous | 0.73 (0.55–0.98) | 0.85 (0.62–1.15) |
| Ethnicity: Other | 0.89 (0.70–1.13) | 1.00 (0.78–1.28) |
| Household income quintile: Q5 (highest; reference) | 1.00 (ref) | 1.00 (ref) |
| Household income quintile: Q1 (poorest) | 1.61 (1.31–1.98) | 1.55 (1.23–1.96) |
| Household income quintile: Q2 | 1.25 (1.02–1.53) | 1.31 (1.05–1.63) |
| Household income quintile: Q3 | 1.03 (0.84–1.25) | 1.08 (0.88–1.34) |
| Household income quintile: Q4 | 0.60 (0.49–0.72) | 0.61 (0.50–0.75) |
| Area of residence: Urban (reference) | 1.00 (ref) | 1.00 (ref) |
| Area of residence: Rural | 0.73 (0.63–0.85) | 0.67 (0.57–0.80) |

In an adjusted median (quantile) regression model (τ = 0.50), non-standardized access remained associated with shorter waiting time, with an adjusted median difference of −3.2 days (95% CI −4.6 to −1.8) compared with standardized access.

Among direct-access specialties (Dentistry, General Medicine, and Psychology), waiting time differed markedly by access type (table 3). Participants with standardized access had a median waiting time of 30 days (IQR 15–30), whereas those reporting non-standardized access had a median of 15 days (IQR 5–30), corresponding to a 15-day shorter median waiting time for non-standardized access (median difference −15·0 days; 95% CI −15·0 to −6·0). In contrast, for referral-required specialties, waiting times were more similar between access types: the median was 30 days (IQR 15–45) for standardized access and 25 days (IQR 15–45) for non-standardized access, yielding a smaller difference of 5 days (median difference −5·0 days; 95% CI −5·0 to 0·0). Overall, the difference in waiting time between access pathways was substantially larger for specialties that did not require referral in routine practice.

**Table 3:** Waiting time to specialist appointments by access type, stratified by referral requirement of the specialty. Ecuador, 2025.

| Specialty stratum | Standard access, n | Standard waiting time, median (IQR), days | Non-standard access, n | Non-standard waiting time, median (IQR), days | Median difference (non – standard), days (95% CI)† |
| --- | --- | --- | --- | --- | --- |
| **Direct-access specialties (Dentistry, General medicine, Family Medicine Psychology) \*** | 130 | 30 (15–30) | 281 | 15 (5–30) | $-15.0$ ($-15.0$ to $-6.0$) |
| **Referral-required specialties (all other specialties)** | 1301 | 30 (15–45) | 2724 | 25 (15–45) | $-5.0$ ($-5.0$ to $0.0$) |

\* The direct access specialties were defined using the specialty codes Dentistry, General Medicine, Family Medicine and Psychology.
†Median differences and 95% CIs were estimated using bootstrap resampling.

## Discussion

This study found that, among adults who attended a scheduled outpatient specialist consultation in Ecuadorian public hospitals, those who reported non-standardized access had shorter waiting times than those who reported standardized access. The difference in waiting time was larger in direct-access specialties than in referral-required specialties. Taken together, these findings suggest that, within routine outpatient specialist care, parallel access pathways may shape the timeliness of care.

Existing evidence suggests that waiting time is a common non-price rationing mechanism in publicly financed systems, but it is not necessarily experienced equally across social groups [3,4,7]. Studies using survey data from European settings have shown that higher socioeconomic position, particularly education, can be associated with shorter waits for specialist consultation, challenging the assumption that waiting lists are inherently equitable [4,6]. More recent cross-country analyses likewise indicate that socioeconomic gradients in waiting time can persist even in systems designed to provide universal coverage, reflecting differences in patients’ ability to navigate care pathways and in how resources and capacity constraints are managed [3,13]. Although much of this literature comes from OECD contexts, qualitative and mixed evidence from Latin America has highlighted that long waiting times after referral are a recurrent organizational barrier within public care networks, often interacting with geographic access barriers and unequal support networks [8,14].

Our findings complement this literature by explicitly quantifying a parallel phenomenon that is frequently discussed but little measured in individual-level studies: the coexistence of standardized and non-standardized access pathways to specialist care, and their relationship to waiting time. Evidence from a recent systematic review on self-referral and direct-access pathways suggests that alternative access routes can have mixed effects but may frequently widen inequalities, because individuals with greater resources or social advantage are more likely to use them effectively [15]. In this context, the shorter waiting times observed among those using non-standardized access are consistent with the idea that non-standardized pathways may bypass bottlenecks in standard scheduling processes, but they also raise concerns that timeliness may depend partly on social connections rather than clinical need, particularly in settings where institutional capacity and referral governance are constrained.

Several mechanisms could plausibly explain why non-standardized access was associated with shorter waiting times in our context. First, non-standardized access may reflect the mobilization of “connection” social capital, whereby personal links with individuals with institutional influence provide privileged information (e.g., on appointment availability) or facilitate overcoming administrative hurdles that would otherwise slow down the standard process [16]. In publicly funded systems with limited specialist capacity, these advantages may translate into earlier appointment allocation without necessarily implying a change in the underlying clinical need.

Second, the observed pattern is compatible with bypassing behaviour documented in other health systems, in which patients do not follow the intended gatekeeping sequence and instead seek direct entry to higher-level care, often influenced by perceived quality, convenience, or prior experience with primary care and its coordination function [17]. Even where standard rules exist, weak enforcement or parallel channels can enable direct entry, particularly when patients perceive the standard route as slow or uncertain.

Third, our stratified findings suggest that institutional norms regarding referral requirements can influence the extent to which non-standardized mechanisms can expedite access. When referral requirements are not binding in practice for direct-access specialties, non-standardized pathways can more easily influence appointment scheduling and are associated with shorter wait times.

And, when it comes to specialties that require referrals, the inequalities in wait times are greater, with those accessing services through standardized pathways having to wait longer. This could pose a risk to the condition of patients waiting through standardized pathways because their need is not self-perceived; it was a physician, specialist or general practitioner, who referred them to the service they are seeking. In contrast, individuals accessing services through non-standardized pathways may arrive at the specialty service based on a self-perception of their condition, which is very likely inaccurate. That is, a headache does not always require examination by a neurologist. For these types of situations, it is necessary to go to the primary care level of the health system, and only when the technical and structural capacities of that center are insufficient to resolve the patient’s health problem does the professional refer them to a more complex hospital [11]. More generally, evidence from diverse settings indicates that social networks and perceived support can affect the use of healthcare and the choice of centers, especially where geographical barriers and access frictions are significant. This may align with our rationale for considering travel time and transportation costs as contextual indicators of access limitations [16].

The distribution of access pathways and waiting times in our study also points to meaningful heterogeneity that is relevant to equity in the Ecuadorian public system. We observed that the likelihood of using non-standardized access varied across sociodemographic strata, including differences by sex, area of residence, geographic region, and income quintile. These patterns are consistent with the broader evidence that health system experiences in Latin America, including “reasonable waiting times”, can display socioeconomic gradients that favor people with higher income and education [18]. At the same time, our results suggest that non-standardized pathways are not exclusively the domain of socially advantaged groups; rather, they may also reflect coping strategies in contexts where standard scheduling processes are perceived as slow or uncertain, and where individuals rely on social ties to overcome administrative frictions.

The lower propensity to use non-standardized access among rural residents in our sample may be explained by structural constraints in rural settings, including fewer specialist resources, longer travel distances, and weaker institutional connectivity. These factors can reduce opportunities to activate influential networks within the health system. Evidence from other contexts indicates that rural populations often face systematic barriers to specialty care, including provider scarcity and longer travel distances, which can compound disadvantages even when standard coverage exists [19]. Our regional differences further support a territorial interpretation: where specialist supply, administrative capacity, and interfacility referral connectivity differ by macro-region, non-standardized access may emerge differentially as either a substitute for, or a bypass of, the standard route.

The socioeconomic patterning we observed also resonates with the longstanding debate that waiting lists, although often framed as an equitable rationing mechanism, can generate unequal non-monetary prices when individuals differ in their ability to navigate the system, mobilize information, or secure timely appointments through non-standardized channels [4,6]. In this sense, non-standardized access can be understood as one manifestation of social capital in health-care utilization: social ties and perceived support may facilitate access to services, but their distribution can also reproduce inequities if the ability to leverage such ties is socially patterned. Notably, the presence of heterogeneity in both access pathways and waiting times strengthens the policy relevance of our findings, because it suggests that reforms aimed solely at reducing average waiting time may be insufficient unless they also address the mechanisms through which some groups can consistently obtain faster access than others.

Our analytic strategy was designed to align with the distributional characteristics of waiting time and to provide estimates that are interpretable for policy. Waiting time for specialist appointments was strongly right-skewed, so we first used a non-parametric comparison to assess whether the overall distributions differed between standardized and non-standardized access, and we presented this comparison visually to avoid overreliance on a single summary statistic. We also reported a rank-based effect size to complement the hypothesis test.

This study has several limitations. First, the analytic sample included only individuals who successfully attended a scheduled outpatient specialist consultation. Therefore, the study does not capture people who sought specialist care but did not obtain an appointment, abandoned the process before attendance, or sought care outside the public system. Second, waiting time and access pathway were self-reported and may be affected by recall error or under-reporting, particularly for non-standardized access. Third, although emergency care and unscheduled urgent visits were excluded by design, we did not directly measure clinical priority among scheduled outpatient consultations, so residual confounding by consultation priority remains possible.

Fourth, the cross-sectional design limits causal interpretation. Fifth, the study was hospital-based and restricted to public hospitals, which may limit generalizability beyond participating public-sector settings. Sixth, scheduling practices and specialist availability may vary across hospitals, and we did not explicitly model hospital-level heterogeneity. Finally, analyses were based on complete cases for each model, which may have introduced bias if missingness was not random.

These findings suggest that reducing waiting times in outpatient specialist care may require not only additional specialist capacity but also greater transparency in how appointments are allocated. Among patients who successfully reached specialist care, shorter waiting times through non-standardized access pathways indicate that timeliness may depend partly on factors outside the formal scheduling process. This pattern appears particularly relevant in specialties where referral requirements are weaker in routine practice. Future research should extend beyond those who attended scheduled consultations in order to capture unmet demand, failed attempts to obtain appointments, and movement between public and private care.

## Data Availability

The data used for our results is available at: https://drive.google.com/file/d/1ny6wF_bndQ2-tCkoE3mJ0uS2tgInZlJ-/view?usp=drive_link Due to its size, it was not possible to share it in supplementary files.

## Funding

This study received no external funding and was supported by Universidad de Especialidades Espíritu Santo (UEES), which had no role in the study or publication process.

## Acknowledgements

We would like to thank the Research Center of the Universidad Espíritu Santo for their trust and support in the creation of this study. We thank all the interviewers who were outside public hospitals collecting the information necessary for this publication. We also thank Ms. Aracely Briones Cevallos for her support of the principal investigator of this project.

## Notes

### Competing Interest Statement

The authors have declared no competing interest.

### Funding Statement

The author(s) received no specific funding for this work.

## References

1. Kruk ME, Gage AD, Arsenault C, Jordan K, Leslie HH, Roder-DeWan S, et al. High-quality health systems in the Sustainable Development Goals era: time for a revolution. Lancet Glob Health. 2018;6: e1196–e1252. 10.1016/s2214-109x(18)30386-3

2. Levesque JF, Harris MF, Russell G. Patient-centred access to health care: conceptualising access at the interface of health systems and populations. Int J Equity Health. 2013;12. 10.1186/1475-9276-12-18

3. Landi S, Ivaldi E, Testi A. Socioeconomic status and waiting times for health services: An international literature review and evidence from the Italian National Health System. Health Policy (New York). 2018;122: 334–351. 10.1016/j.healthpol.2018.01.003

4. Laudicella M, Siciliani L, Cookson R. Waiting times and socioeconomic status: Evidence from England. Soc Sci Med. 2012;74: 1331–1341. 10.1016/j.socscimed.2011.12.049

5. Sharma A, Siciliani L, Harris A. Waiting times and socioeconomic status: Does sample selection matter? Econ Model. 2013;33: 659–667. 10.1016/j.econmod.2013.05.009

6. Siciliani L, Verzulli R. Waiting times and socioeconomic status among elderly europeans: Evidence from share. Health Econ. 2009;18: 1295–1306. 10.1002/hec.1429

7. Johar M, Jones G, Keane MP, Savage E, Stavrunova O. Discrimination in a universal health system: Explaining socioeconomic waiting time gaps. J Health Econ. 2013;32: 181–194. 10.1016/j.jhealeco.2012.09.004

8. Vargas I, Garcia-Subirats I, Mogollón-Pérez AS, Ferreira-De-Medeiros-Mendes M, Eguiguren P, Cisneros AI, et al. Understanding communication breakdown in the outpatient referral process in Latin America: a cross-sectional study on the use of clinical correspondence in public healthcare networks of six countries. Health Policy Plan. 2018;33: 494–504. 10.1093/heapol/czy016

9. Herrera CA, Bascolo E, Villar-Uribe M, Houghton N, Bennett S, Castro MC, et al. No time to wait: resilience as a cornerstone for primary health care across Latin America and the Caribbean, a World Bank-PAHO Lancet Regional Health Americas Commission. The Lancet Regional Health – Americas. 2025;50: 101240. 10.1016/j.lana.2025.101240

10. Penchansky R, Thomas JW. The concept of access: definition and relationship to consumer satisfaction. Med Care. 1981;19: 127–140. 10.1097/00005650-198102000-00001

11. Ministerio de Salud Pública. Modelo de Atención Integral del Sistema Nacional de Salud. Quito; 2013. Available: http://instituciones.msp.gob.ec/images/Documentos/Ministerio/sub_gobernanza_salud/manual_mais_2013.pdf

12. Armijos Briones M, Figueroa Intriago S, Lanata-Flores A, Benitez Sellán P, Marcillo Toala O, Ayala Aguirre PE. Protocol: Waiting time and ways of accessing specialized health services in public hospitals in Ecuador. PLoS One. 2025;20: e0315149. 10.1371/journal.pone.0315149

13. Martin S, Siciliani L, Smith P. Socioeconomic inequalities in waiting times for primary care across ten OECD countries. Soc Sci Med. 2020;263: 113230. 10.1016/j.socscimed.2020.113230

14. Vargas I, Mogollón-Pérez AS, Eguiguren P, Torres AL, Peralta A, Rubio-Valera M, et al. Understanding the health system drivers of delayed cancer diagnosis in public healthcare networks of Chile, Colombia and Ecuador: A qualitative study with health professionals, managers and policymakers. Soc Sci Med. 2025;365: 117499. 10.1016/j.socscimed.2024.117499

15. Harvey-Sullivan A, Lynch H, Tolley A, Gitlin-Leigh G, Kuhn I, Ford JA. What impact do self-referral and direct access pathways for patients have on health inequalities? Health Policy (New York). 2024;139: 104951. 10.1016/j.healthpol.2023.104951

16. Pitkin Derose K, Varda DM. Social Capital and Health Care Access. Medical Care Research and Review. 2009;66: 272–306. 10.1177/1077558708330428

17. Aoki T, Yamamoto Y, Ikenoue T, Kaneko M, Kise M, Fujinuma Y, et al. Effect of Patient Experience on Bypassing a Primary Care Gatekeeper: a Multicenter Prospective Cohort Study in Japan. J Gen Intern Med. 2018;33: 722. 10.1007/s11606-017-4245-1

18. Roberti J, Leslie HH, Doubova S V., Ranilla JM, Mazzoni A, Espinoza L, et al. Inequalities in health system coverage and quality: a cross-sectional survey of four Latin American countries. Lancet Glob Health. 2024;12: e145–e155. 10.1016/s2214-109x(23)00488-6

19. Herberholz C, Phuntsho S. Social capital, outpatient care utilization and choice between different levels of health facilities in rural and urban areas of Bhutan. Soc Sci Med. 2018;211: 102–113. 10.1016/j.socscimed.2018.06.010

